# Prospective Observational Study of Screening Asymptomatic Healthcare Workers for SARS-CoV-2 at a Canadian Tertiary Care Center

**DOI:** 10.1101/2020.07.21.20159053

**Authors:** Deepali Kumar, Victor H. Ferreira, Andrzej Chruscinski, Vathany Kulasingam, Trevor J. Pugh, Tamara Dus, Brad Wouters, Amit Oza, Matthew Ierullo, Terrance Ku, Beata Majchrzak-Kita, Sonika T. Humar, Ilona Bahinskaya, Natalia Pinzon, Jianhua Zhang, Lawrence E. Heisler, Paul M. Krzyzanowski, Bernard Lam, Ilinca M. Lungu, Dorin Manase, Krista M. Pace, Pouria Mashouri, Michael Brudno, Michael Garrels, Tony Mazzulli, Myron Cybulsky, Atul Humar

**Affiliations:** University Health Network, Toronto, Ontario, Canada; Sinai Health System, Toronto, Ontario, Canada; Ontario Institute for Cancer Research, Toronto, Ontario, Canada; University of Toronto, Toronto, Ontario, Canada; University Health Network Digital, Toronto, Ontario, Canada

## Abstract

We screened three separate cohorts of healthcare workers for SARS-CoV-2 via nasopharyngeal swab PCR. A seroprevalence analysis using multiple assays was performed in a subgroup. The asymptomatic health care worker cohorts had a combined swap positivity rate of 29/5776 (0.50%, 95%CI 0.32-0.75) compared to the symptomatic cohort rate of 54/1597 (3.4%) (ratio of symptomatic to asymptomatic 6.8:1). Sequencing demonstrated several variants. The seroprevalence (n=996) was 1.4-3.4% depending on assay. Protein microarray analysis showed differing SARS-CoV-2 protein reactivities and helped define likely true positives vs. suspected false positives. Routine screening of asymptomatic health care workers helps identify a significant proportion of infections.

## BACKGROUND

SARS-CoV-2 is a novel respiratory coronavirus that has evolved into a widespread global pandemic. The transmission of COVID-19 to healthcare workers (HCWs) from patients, colleagues, or the community is a serious concern as it places potentially highly vulnerable patient populations at risk. Symptom screening for HCWs is standard infection control practice to help mitigate spread and protect both patients and other HCWs. However, studies have shown that a significant proportion of individuals have asymptomatic or pre-symptomatic infection but may still transmit virus ^1-5^. We sought to understand the prevalence of asymptomatic SARS-CoV-2 infection in HCWs in a large Canadian tertiary care center (including a referral center for severe COVID-19) to determine the potential benefits of asymptomatic HCW screening. We also determined the seroprevalence of SARS-CoV-2 antibodies in HCW using commercial and in-house assays.

## METHODS

The setting for the study was the University Health Network, a large tertiary care center in Toronto, Canada with multiple sites and approximately 1300 total inpatient beds. The center includes both acute and long-term facilities and a provincial referral unit for advanced lung support for COVID-19 patients, as well as several dedicated COVID units. Over a 6 week period, HCWs were prospectively enrolled and underwent between 1-6 serial nasopharyngeal swabs for SARS-CoV-2 polymerase chain reaction (PCR) testing with communication and action in response to results in real-time. HCW were required to be asymptomatic and not have a previous diagnosis of COVID. Any symptomatic health care worker was referred to hospital occupational health and safety. The study was approved by the institutional research ethics board. Additional HCWs (asymptomatic or symptomatic) self-identified for voluntary screening through occupational health and safety. During the study period, the hospital cared for 975 COVID-19 patients of which approximately one-third were inpatients. Universal masking was in effect in the hospital and HCW with direct patient contact were required to wear a face shield. N95 masks were reserved for aerosol generating procedures.

### SARS-CoV-2 PCR

Nasopharyngeal swabs were collected and underwent PCR testing by the clinical microbiology laboratory using either the Seegene Allplex PCR assay (Korea) or Altona PCR assay (Altona Diagnostics, Germany) using manufacturer’s instructions.

### Serology testing

Serologic testing for anti-SARS-CoV-2 IgG antibody was performed on a subset of consenting individuals. Serology was performed using two commercially available IgG assays, one that tests anti-nucleoprotein (NP) antibodies by CMIA (Abbott Diagnostics, Health Canada approved assay) and the other for anti-spike (S) antibodies (EuroImmun, Germany) followed by a further assessment using a custom in-house protein microarray platform. Commercial assays were carried out using manufacturer’s instructions. To confirm antibody specificities a custom microarray was performed using 45 commercially available coronavirus recombinant proteins corresponding to SARS-CoV-2, SARS-CoV, MERS-CoV and community coronaviruses (CoV-NL63, -HKU1, - 229E and -OC43) (Sino Biological and ProSci) ^6,7^. [See Supplementary methods and Supplementary Table 1].

### Viral genome sequencing

Targeted sequencing of the SARS-CoV-2 genome was performed for nasopharyngeal swab samples that were positive by PCR (See Supplementary methods). Briefly, RNA was isolated from nasopharyngeal swab fluid, followed by RT-PCR and PCR amplification of the complete viral genome using the ARTIC network version 3 primer set ^8^. PCR products were sequenced on the Illumina MiSeq system using V2 sequencing chemistry and 250 bp paired end reads. Reads were aligned to the SARS-CoV-2 reference genome (GenBank: MN908947.3) using a Nextflow workflow ^9^ to form consensus sequences using the ARTIC network nCoV-2019 novel coronavirus bioinformatics protocol^10^. Only samples having >75% of the SARS-CoV-2 genome with consensus calls were used.

## RESULTS

Three separate cohorts were analyzed (Figure 1). The primary study cohort was a total of 1669 HCWs that were enrolled over a 6-week period (April 17 – May 29, 2020) with a total of 3173 nasopharyngeal swabs performed. HCWs included primarily nurses (n=655), physicians (n=152), allied health (n=446) and other (Supplementary Table 2). Absence of symptoms was confirmed for all participants at the time of testing.

**FIGURE 1A:**
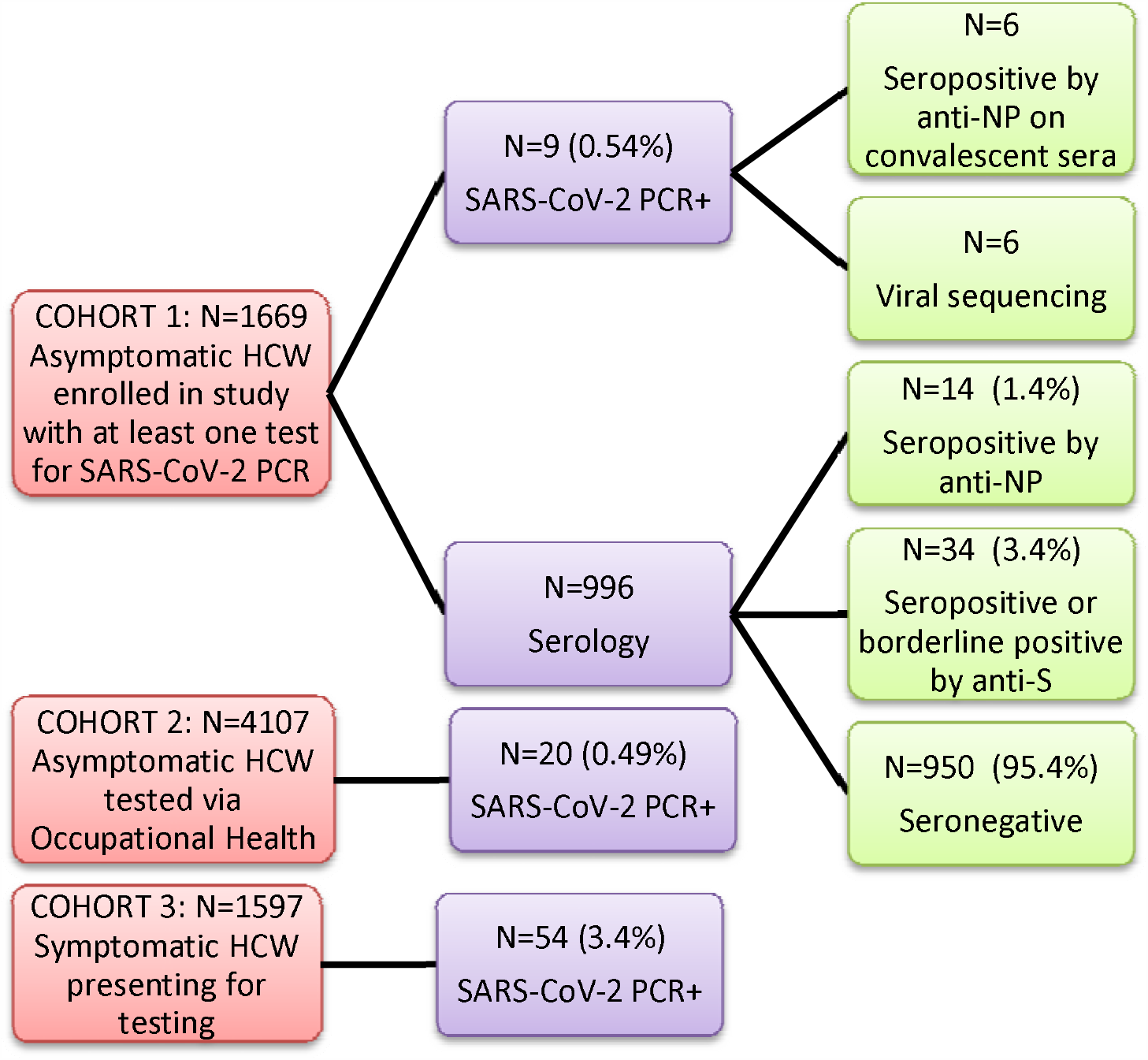
Study Flow and Outcomes. HCW healthcare workers, NP nucleoprotein, S spike.

472/1555 (29.1%) were actively involved in the care of COVID patients in the immediate two weeks prior to at least one of their swabs. The second cohort consisted of an additional 4107 asymptomatic HCWs who were tested through voluntary testing in the occupational health and safety department. The third cohort was symptomatic and consisted of an additional 1597 HCWs who self-identified as having at least one symptom compatible with COVID-19 and were tested by nasopharyngeal swab for SARS-CoV-2.

### SARS-CoV-2 PCR

The prevalence of a positive nasopharyngeal swab at any time point in the primary study asymptomatic cohort was 9/1669 (0.54%, 95% CI 0.28-1.02). Of the 9 positive HCWs, 4 subsequently developed symptoms while the rest remained asymptomatic/pauci-symptomatic (Supplementary Table 3). Follow-up investigations in the patient wards of positive HCWs helped identify and subsequently contain two separate outbreaks in which previously unidentified patients were also found to be positive (data not shown). In the secondary volunteer cohort of 4107 asymptomatic HCWs, 20 were positive (0.49%, 95%CI 0.32-0.75) for a combined swab positive prevalence of 29/5776 (0.50%, 95%CI 0.35-0.72) in the two cohorts. In the self-identified HCW cohort with symptoms during the same 6-week period, 54/1597 (3.4%) were found to be PCR positive. Based on this, the ratio of symptomatic to asymptomatic positive HCWs was approximately 6.8 to 1.

### Sequencing

Sequencing was successful in 6/9 positive HCWs in cohort 1. Based on sequence analysis, three predominant viral strains were identified (Supplementary Figure 1). In conjunction with an analysis of ward locations for positive HCWs, this suggests that at least some of the positive cases may have been due to HCW to HCW transmission or possibly from a common patient source.

### Serology testing

A subset of 996 HCWs also underwent serology testing (Supplementary Table 2). By the anti-NP CMIA serology assay, a total of 14/996 (1.4%) were IgG positive (Supplementary Table 4). By the anti-Spike assay, a total of 22/996 HCWs were IgG positive (2.2%) and an additional 12/996 (1.2%) had borderline positive results.

However, only two HCWs were IgG positive in both assays. We then analyzed all 34 seropositives via protein microarray to confirm antibodies against specific SARS CoV-2 proteins (Figure 2A-B). In the 14 HCWs positive by anti-NP, 13/14 had evidence of IgG antibodies against SARS CoV-2 nucleoprotein and 5 had evidence of antibodies against other viral proteins including spike protein and its receptor binding domain. Of the 22 positive by anti-Spike ELISA, 5 had evidence of antibodies against at least one SARS-CoV-2 spike protein and only the two that were positive by both assays had evidence of antibodies against nucleoprotein.

**Figure 2.**
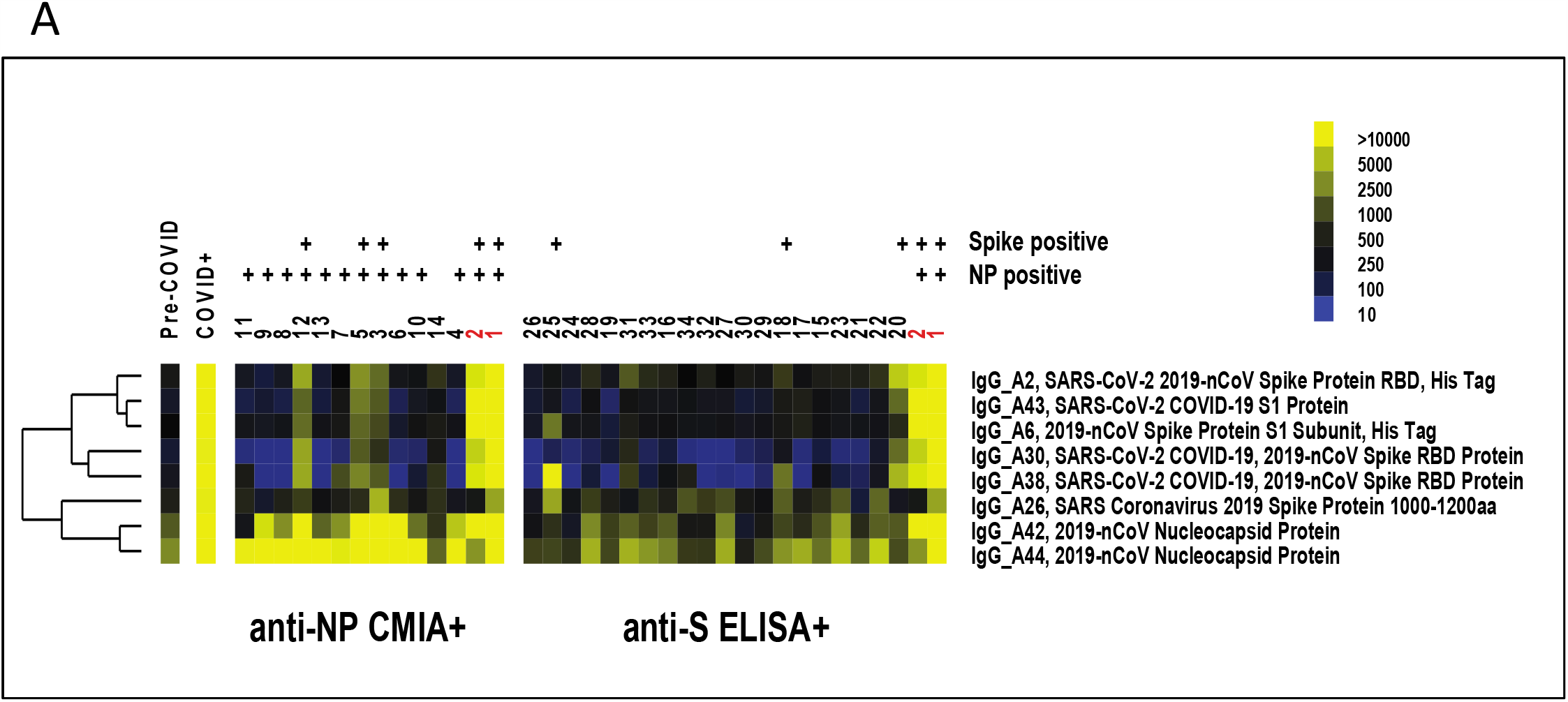
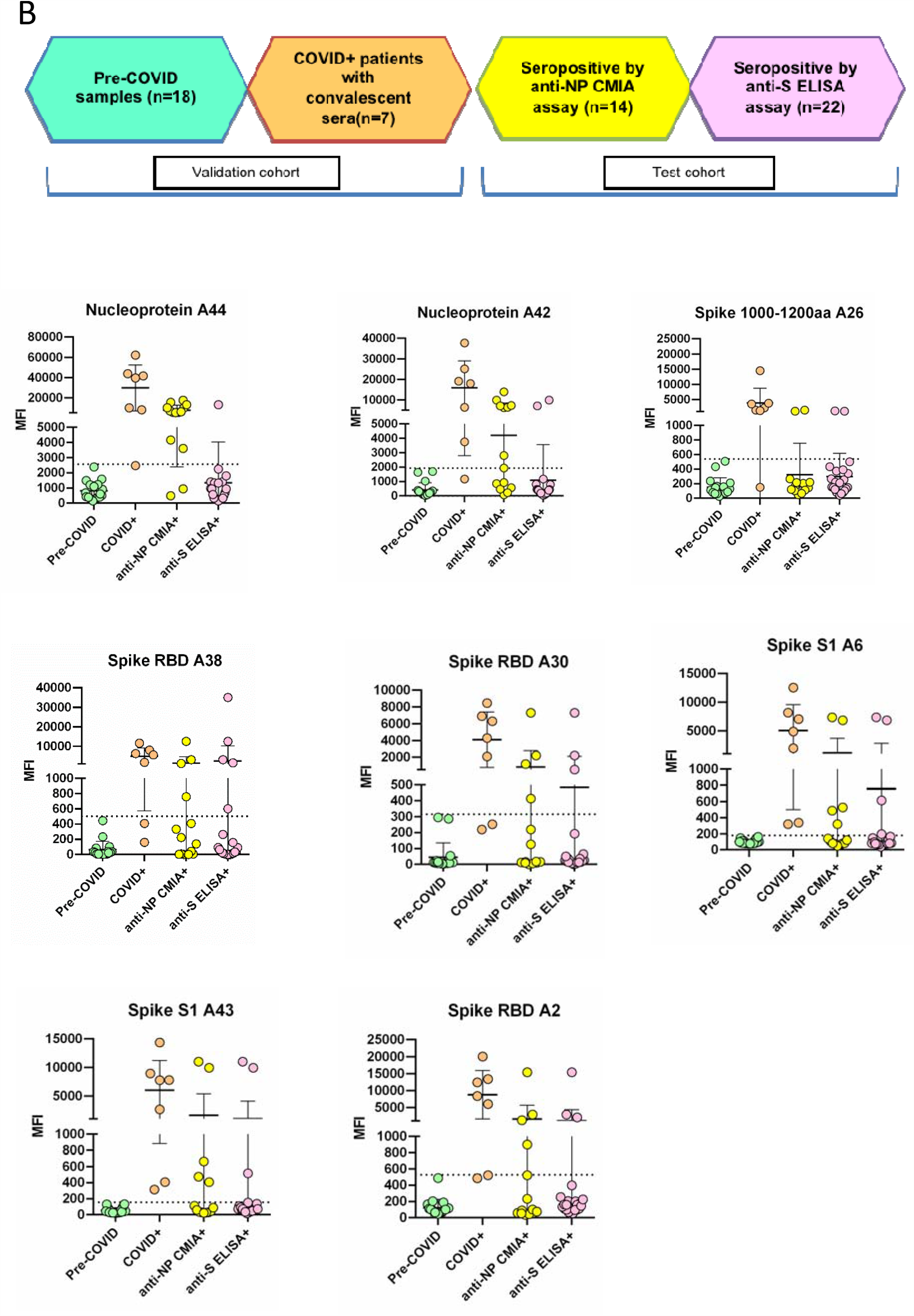
Heatmap and graph of SARS-CoV-2 reactivity in study samples on antigen microarrays. A)Heatmap of eight SARS-CoV-2 IgG reactivities in individual anti-NP CMIA+ and anti-S ELISA+ samples. These antigen reactivities represent the eight highest ranked IgG reactivities that are upregulated in COVID+ samples (with mean MFI-B > 1000) as determined by significance analysis of microarrays. Anti-NP and anti-spike reactivity in individual samples on the arrays is indicated above the sample numbers. Positivity on the arrays was determined as described below. The mean reactivity of pre-COVID and COVID+ samples is shown as a comparison. The sample numbers in red indicate dual positive (anti-NP CMIA+ and anti-S ELISA+) samples. Yellow indicates high reactivity, whereas blue indicates low reactivity on the heatmap. B) Graphs of individual antigen IgG reactivity (MFI-B) in pre-COVID, COVID+, anti-NP CMIA+ and anti-S ELISA+ groups. Graphs show mean ∓ SD for samples in each of the groups. Samples in the anti-NP CMIA+ and anti-S ELISA+ groups were considered positive if the MFI-B was higher than the mean + 3 SD of the pre-COVID samples (dotted line). MFI-B, median fluorescent intensity minus background; NP, nucleocapsid protein; S, spike; SD, standard deviation.

## DISCUSSION

This study demonstrates that routine SARS-CoV-2 PCR screening of asymptomatic HCWs in a large tertiary care hospital was valuable to identify and act upon unrecognized SARS-CoV-2 infection. Some of the positive HCWs were pre-symptomatic while others remained asymptomatic throughout their clinical course. We also found that in the hospital setting, there were significant numbers of asymptomatic infections with the ratio of symptomatic to asymptomatic HCWs being approximately 6.8:1. Serology demonstrated a higher rate of positivity suggesting that additional sequential PCR screening over time would likely be useful.

Previous studies of HCWs have shown mixed results with regards to symptomatic and asymptomatic infections. In Seattle, Washington, 185/3477 (5.3%) of symptomatic HCWs were found to have nucleic acid positivity for SARS-CoV-2^11^. Hunter et al. screened 1654 HCWs in England and found a 14% rate of positivity with similar rates in non-clinical staff vs. clinical staff^12^. However, no data on symptoms was available in this study. In terms of asymptomatic infection, Lai et al. tested 335 asymptomatic HCWs in Wuhan, China and found 3 positives (0.9%) ^5^. No serologic testing was performed in either study. Our study showed that although our overall active infections were low, a significant proportion were asymptomatic/pre-symptomatic.

In our study, depending on the commercial assay 1.4%-3.4% of HCW had evidence of past SARS-CoV-2 infection. This is similar to a German cohort where the seroprevalence among 406 clinic staff was found to be 2.7% ^13^, but lower than a Spanish cohort of HCWs where 9.3% were seropositive ^14^. A novel aspect of our study was confirmatory assessment using a microarray-based assay to determine protein specific SARS-CoV-2 IgG antibodies which showed varying protein reactivity in HCWs who were seropositive based on commercially available assays. These data coupled with lack of agreement between commercial assays highlight the pitfalls and variability of results when performing large scale serosurveys in lower prevalence asymptomatic populations. In addition, the antibody profile post-infection may differ in individuals depending on clinical course and assays to look at multiple antigens simultaneously provide more robust information. While single-target assays may perform relatively well in patients with known COVID, when applied to large seroprevalence studies, performance characteristics appear poorer with significant disagreement between tests. Assays vary in their protein targets and since antibodies may wane over time, distinguishing true from false positives may be difficult. We suspect several of the results on a single assay were likely false positives.

In summary we show that a significant proportion of HCW during the pandemic may be asymptomatic/pre-symptomatic and propose that if symptomatic HCW in an institution are being diagnosed with COVID, then asymptomatic HCW testing should also be offered. Data on serosurveys in the asymptomatic HCW population need to be carefully interpreted as performance characteristics of assays may vary. However, the generally higher rate of past infections compared to current infections suggests there is utility in sequential screening of asymptomatic HCW by nasopharyngeal swabs.

## Data Availability

Study data are available upon request.

## FUNDING

The study was funded by a peer-reviewed grant from the Mount Sinai Hospital and University Health Network Academic Medical Organization and the Toronto General and Western Hospital Foundation. This study was conducted with the support of the Genomics (genomics.oicr.on.ca) and Diagnostic Development programs of the Ontario Institute for Cancer Research through funding provided by the Government of Ontario.

## AUTHOR CONTRIBUTIONS

D.K., A.H., B.W., A.O. developed the study concept and protocol. D.K., V.H.F., A.C., V.K., T.J.P., L.E.H., P.M.K., B.L., I.M.L., T.D., M.I., T.K, B.M-K., J.Z., I.B, N.P, M.G., T.M., M.C., D.M., K.M.P., P.M., M.B. performed the research, analyzed, and collected data. All authors contributed to the writing of the manuscript.

## COMPETING INTERESTS

None for all authors.

## SUPPLEMENTARY LABORATORY METHODS

### Viral genome sequencing

Targeted sequencing of the SARS-CoV-2 genome was performed for nasopharyngeal swab samples that were positive by PCR. Briefly, RNA was isolated from nasopharyngeal swab fluid using Mag-Bind Viral DNA/RNA 96 Kit, and RT-PCR was performed using SuperScript IV First Strand Synthesis System (Thermo Fisher) and Q5 Hot Start High-Fidelity DNA Polymerase (NEB). The complete viral genome was amplified using a set of overlapping PCR primers, version 3 developed by the ARTIC network ^8^. PCR products were sequenced on an Illumina MiSeq system using 250 bp paired end reads. Read were aligned to the SARS-CoV-2 reference genome (GenBank: MN908947.3) using a Nextflow workflow ^9^ that generates a consensus sequence from Illumina reads using the ARTIC network nCoV-2019 novel coronavirus bioinformatics protocol ^10^. Consensus calls required a minimum coverage depth of 10, with a frequency threshold of 0.75 to call a variant. Only samples with >75% of the SARS-CoV-2 genome having consensus calls were used.

### Antibody Testing

10mL of peripheral blood was collected in red-top tubes (BD Vacutainer, Fisher Scientific, Mississauga, ON, Canada), incubated for at least 30 minutes to allow for clotting, and subsequently centrifuged at 2000 RCF for 10 minutes. Serum was collected in cryovials and frozen at -80 for batch processing. The EUROIMMUN anti-SARS-CoV-2 ELISA (IgG) kit ^15^ (EUROIMMUN AG, Luebeck, Germany) was performed manually. Briefly, serum was thawed and diluted 1:101 and added to wells pre-coated with antigens corresponding to the S1 region of the spike protein. To detect the bound antibodies, a second incubation is carried out using an enzyme-labelled anti-human IgG and substrate catalyzing a colorimetric reaction. Results are evaluated semi-quantitatively by calculation of a ratio of the extinction of the control or patient sample over the extinction of the calibrator. This ratio is interpreted as follows: < 0.8 negative, ≥0.8 to <1.0 borderline, and ≥ 1.1 IgG positive. The Abbott SARS-CoV-2 IgG ^16^ assay is a chemiluminescent microparticle immunoassay (CMIA) run on the fully automated ARCHITECT instrument (Abbott Laboratories, Chicago, IL, USA). Briefly, 75uL of undiluted serum per sample was loaded onto SARS-CoV-2 nucleoprotein coated paramagnetic microparticles, and assay diluent are combined and incubated. After washing, an anti-human IgG acridinium-labeled conjugate is added and the resulting chemiluminescent reaction is measured in relative light units (RLUs). The presence or absence of IgG antibodies to SARS-CoV-2 in the sample was determined by comparing the chemiluminescent RLU in the reaction to the calibrator RLU. An index measurement≥1.4 was considered positive for anti-SARS-CoV-2 IgG antibodies. Both antibody tests received Emergency Use Authorization from the US Food and Drug Administration (FDA); the Abbott test has also received Health Canada authorization.

### Antigen Microarray

The Coronavirus antigen microarray was generated using previously published protocols for generation of antigen microarrays to screen for autoantibodies in heart failure and transplantation ^6,7^. Human IgA, IgM, IgG and viral antigens were spotted in triplicate onto two-pad FAST nitrocellulose-coated slides (GVS North America, Sanford, ME, USA) using a Chipwriter Pro microarrayer (Virtek, Waterloo, ON., Canada) with solid pins (Arrayit, Sunnyvale, CA, USA). Dried slides were placed in FAST frames (GVS North America, Sanford, ME, USA) and blocked overnight at 4°C (blocking buffer: PBS, 5% FBS, 0.1% Tween). The next day, arrays were incubated with patient serum (diluted 1:100 in blocking buffer) for one hour at 4°C. After washing, the slides were incubated for 45 minutes at 4°C with a pair of secondary antibodies consisting of Cy3-labeled goat anti-human IgG (Jackson ImmunoResearch, West Grove, PA, USA) and Alexa Fluor 647-labeled goat anti-human IgM (Jackson ImmunoResearch, West Grove, PA, USA). After drying, fluorescent intensities of features were quantified using an Axon 4200A microarray scanner (Molecular Devices, Sunnyvale, CA., USA) with Genepix 6.1 software (Molecular Devices). Median fluorescent intensity minus local background (MFI-B) was determined at 532nm for Cy3, and 635nm for Alexa Fluor 647. The single averaged MFI-B for each antigen was calculated from the features arrayed in triplicate. A diverse collection of Coronavirus antigens (n=45), targeting SARS-CoV-2, along with MERS-CoV, SARS-CoV and community coronaviruses were used. The antigen library consisted of 30 SARS-CoV-2 antigens, 5 MERS-CoV antigens, 4 SARS-CoV antigens and 6 community coronavirus antigens (Supplementary Table 1). Antigens were diluted to 0.25 mg/ml in PBS and stored in aliquots at -80°C until the day of microarray printing.

In order to validate the results, convalescent sera from known COVID+ persons (n=7) obtained 6 weeks after infection and banked sera from healthy controls obtained prior to COVID-19 (n=18) were tested on the microarray platform. Significance of microarrays (SAM) demonstrated 39 reactivities that were higher in the COVID+ sera compared with pre-COVID samples (Supplementary Figure 3, Supplementary Table 5).

The eight highest ranked IgG reactivities by SAM (with mean MFI-B > 1,000 in COVID+ samples) were used for analysis of study samples that were positive by the two commercial kits (anti-NP CMIA and anti-S ELISA). Images of arrays probed with secondary antibodies only, pre-COVID serum, and COVID+ serum are shown in Supplementary Figure 2A. The linearity of the array assay for antibody detection was demonstrated by probing arrays with serial dilutions of serum from a COVID+ person (Supplementary Figure 2B-C).

**Supplementary Table 1:** Viral Antigens included in Protein Microarray.

| Antigen # | Antigen | Virus | Source | Catalog # | Expression System |
| --- | --- | --- | --- | --- | --- |
| A1 | Human coronavirus (HCoV-229E) Spike Protein (S1 Subunit, His Tag) | Community Coronavirus | Sino Biological | 40601-V08H | HEK293 |
| A2 | SARS-CoV-2 (2019-nCoV) Spike Protein (RBD, His Tag) | SARS-CoV-2 | Sino Biological | 40592-V08H | HEK293 |
| A3 | 2019-nCoV Spike Protein (S1+S2 ECD, His tag) | SARS-CoV-2 | Sino Biological | 40589-V08B1 | Baculovirus-Insect Cells |
| A4 | 2019-nCoV Nucleocapsid Protein (His tag) | SARS-CoV-2 | Sino Biological | 40588-V08B | Baculovirus-Insect Cells |
| A5 | 2019-nCoV Spike Protein (S2 ECD, His tag) | SARS-CoV-2 | Sino Biological | 40590-V08B | Baculovirus-Insect Cells |
| A6 | 2019-nCoV Spike Protein (S1 Subunit, His Tag) | SARS-CoV-2 | Sino Biological | 40591-V08H | HEK293 |
| A7 | SARS-CoV-2/2019-nCoV Plpro / papainlike protease (aa 1564-1880, His Tag) | SARS-CoV-2 | Sino Biological | 40593-V07E | E. Coli |
| A8 | 2019-nCoV Spike Protein (S1 Subunit, His tag) | SARS-CoV-2 | Sino Biological | 40591-V08B1 | Baculovirus-Insect Cells |
| A9 | SARS-CoV-2 (2019-nCoV) Methyltransferase / ME-his Recombinant Protein | SARS-CoV-2 | Sino Biological | 40598-V07E | E. Coli |
| A10 | Human SARS Coronavirus Nucleoprotein / NP Protein (His Tag) | SARS-CoV | Sino Biological | 40143-V08B | Baculovirus-Insect Cells |
| A11 | Human SARS Coronavirus Spike Protein (S1 Subunit, His Tag) | SARS-CoV | Sino Biological | 40150-V08B1 | Baculovirus-Insect Cells |
| A12 | Human SARS Coronavirus Spike Protein (RBD, His Tag) | SARS-CoV | Sino Biological | 40150-V08B2 | Baculovirus-Insect Cells |
| A13 | SARS-CoV (strain WH20) Plpro / papain-like protease (His Tag) | SARS-CoV | Sino Biological | 40524-V08E | E. Coli |
| A14 | MERS-CoV (NCoV / Novel coronavirus) Nucleoprotein / NP protein (His Tag) | MERS-CoV | Sino Biological | 40068-V08B | Baculovirus-Insect Cells |
| A15 | MERS-CoV (NCoV / Novel coronavirus) Spike Protein (S1 Subunit, aa 1-725, His Tag) | MERS-CoV | Sino Biological | 40069-V08H | HEK293 |
| A16 | MERS-CoV (NCoV / Novel coronavirus) Spike Protein (S2 Subunit, aa 726-1296, His Tag) | MERS-CoV | Sino Biological | 40070-V08B | Baculovirus-Insect Cells |
| A17 | MERS-CoV (NCoV / Novel coronavirus) Spike Protein fragment (RBD, aa 367-606, His Tag) | MERS-CoV | Sino Biological | 40071-V08B1 | Baculovirus-Insect Cells |
| A18 | MERS-CoV (NCoV / Novel coronavirus) Spike Protein (S1 Subunit, aa 1-725, His Tag) | MERS-CoV | Sino Biological | 40069-V08B1 | Baculovirus-Insect Cells |
| A19 | Human coronavirus spike glycoprotein Protein (isolate HKU1) (S1 Subunit, aa 1-760, His Tag) | Community Coronavirus | Sino Biological | 40021-V08H | HEK293 |
| A20 | Human coronavirus HKU1 (isolate N5) (HCoV-HKU1) Spike/S1 Protein (S1 Subunit, His Tag) | Community Coronavirus | Sino Biological | 40602-V08H | HEK293 |
| A21 | Human coronavirus (HCoV-229E) Spike Protein (S1+S2 ECD, His Tag) | Community Coronavirus | Sino Biological | 40605-V08B | Baculovirus-Insect Cells |
| A22 | Human coronavirus (HCoV-NL63) Spike/S1 Protein (S1 Subunit, His Tag) | Community Coronavirus | Sino Biological | 40600-V08H | HEK293 |
| A23 | Human coronavirus (HCoV-OC43) Hemagglutinin esterase Protein (His Tag) | Community Coronavirus | Sino Biological | 40603-V08H | HEK293 |
| A24 | MERS-CoV (NCoV / Novel coronavirus) Spike Protein (ECD, aa 1-1297, His Tag) | MERS-CoV | Sino Biological | 40069-V08B | Baculovirus-Insect Cells |
| A25 | SARS Coronavirus 2019 Spike Recombinant protein (800-1000 aa) | SARS-CoV-2 | ProSci Inc | 39-125 | E. Coli |
| A26 | SARS Coronavirus 2019 Spike Recombinant protein (1000-1200 aa) | SARS-CoV-2 | ProSci Inc | 39-126 | E. Coli |
| A27 | SARS Coronavirus 2019 Nucleocapsid Recombinant protein | SARS-CoV-2 | ProSci Inc | 39-113 | E. Coli |
| A28 | SARS Coronavirus 2019 Nucleocapsid Mosaic Recombinant protein | SARS-CoV-2 | ProSci Inc | 39-115 | E. Coli |
| A29 | SARS Coronavirus 2019 Spike E Mosaic Recombinant protein | SARS-CoV-2 | ProSci Inc | 39-114 | E. Coli |
| A30 | SARS-CoV-2 (COVID-19, 2019-nCoV) Spike RBD Recombinant Protein | SARS-CoV-2 | ProSci Inc | 10-100 | Human cells |
| A31 | SARS-CoV-2 (COVID-19, 2019-nCoV) S1+S2 ECD (S-ECD) Recombinant Protein | SARS-CoV-2 | ProSci Inc | 10-108 | Human cells |
| A32 | SARS-CoV-2 (COVID-19, 2019-nCoV) ORF8 Recombinant Protein | SARS-CoV-2 | ProSci Inc | 10-002 | E. Coli |
| A33 | SARS-CoV-2 (COVID-19, 2019-nCoV) ORF3a Recombinant Protein | SARS-CoV-2 | ProSci Inc | 10-005 | E. Coli |
| A34 | SARS-CoV-2 (COVID-19, 2019-nCoV) Spike Recombinant Protein | SARS-CoV-2 | ProSci Inc | 10-006 | E. Coli |
| A35 | SARS-CoV-2 (COVID-19, 2019-nCoV) Nucleocapsid Recombinant Protein | SARS-CoV-2 | ProSci Inc | 10-007 | E. Coli |
| A36 | SARS-CoV-2 (COVID-19, 2019-nCoV) Spike-RBD Recombinant Protein | SARS-CoV-2 | ProSci Inc | 10-008 | Sf21 cells |
| A37 | SARS-CoV-2 (COVID-19, 2019-nCoV) Spike-ECD Recombinant Protein | SARS-CoV-2 | ProSci Inc | 10-011 | Sf21 cells |
| A38 | SARS-CoV-2 (COVID-19, 2019-nCoV) Spike-RBD Recombinant Protein | SARS-CoV-2 | ProSci Inc | 10-015 | Human cells |
| A39 | COVID 19 M Coronavirus Recombinant Protein | SARS-CoV-2 | mybiosource.com | MBS8574735 | E. Coli |
| A40 | SARS-CoV-2 (COVID-19) 3C-like Proteinase | SARS-CoV-2 | ProSci Inc | 10-116 | E. Coli |
| A41 | SARS-CoV-2 (COVID-19) Papain-like Protease | SARS-CoV-2 | ProSci Inc | 10-119 | E. Coli |
| A42 | 2019-nCoV Nucleocapsid Recombinant Protein | SARS-CoV-2 | ProSci Inc | 97-085 | HEK293 |
| A43 | SARS-CoV-2 (COVID-19) S1 Recombinant Protein | SARS-CoV-2 | ProSci Inc | 97-087 | HEK293 |
| A44 | 2019-nCoV Nucleocapsid Recombinant Protein | SARS-CoV-2 | ProSci Inc | 97-077 | E. Coli |
| A45 | 2019-nCoV Envelope Recombinant Protein | SARS-CoV-2 | ProSci Inc | 97-082 | E. Coli |

**Supplementary Table 2:** Characteristics of health care workers undergoing nasopharyngeal swab (in Cohort 1) and serology testing.

| **Variable** | **Nasopharyngeal SARS-CoV-2 PCR**<br>**N=1669** | **Serology**<br>**N=996** |
| --- | --- | --- |
| Age (years); mean ± s.d. | 40.3 ± 11.3 | 40.8 ± 11.1 |
| Sex (M/F/Other) | 356/1312/1 | 215/781 |
| Occupation |  |  |
| Nurse | 655 (39.2%) | 361 (36.2%) |
| Physician | 152 (9.1%) | 101 (10.1%) |
| Allied Health | 446 (26.7%) | 261 (26.2%) |
| Other | 396 (23.7%) | 272 (27.3%) |
| Not specified | 20 (1.2%) | 1 (0.1%) |
| Lives with children <12 years of age | 452/1555 (29.1%) | 266/927 (28.7%) |
| Directly looked after COVID patient in the last 2 weeks | 472/1555 (30.4%) | 255/927 (27.5%) |
| Travel in the last 2 weeks | 2/1555 (0.13%) | 2/927 (0.22%) |

**Supplementary Table 3:**
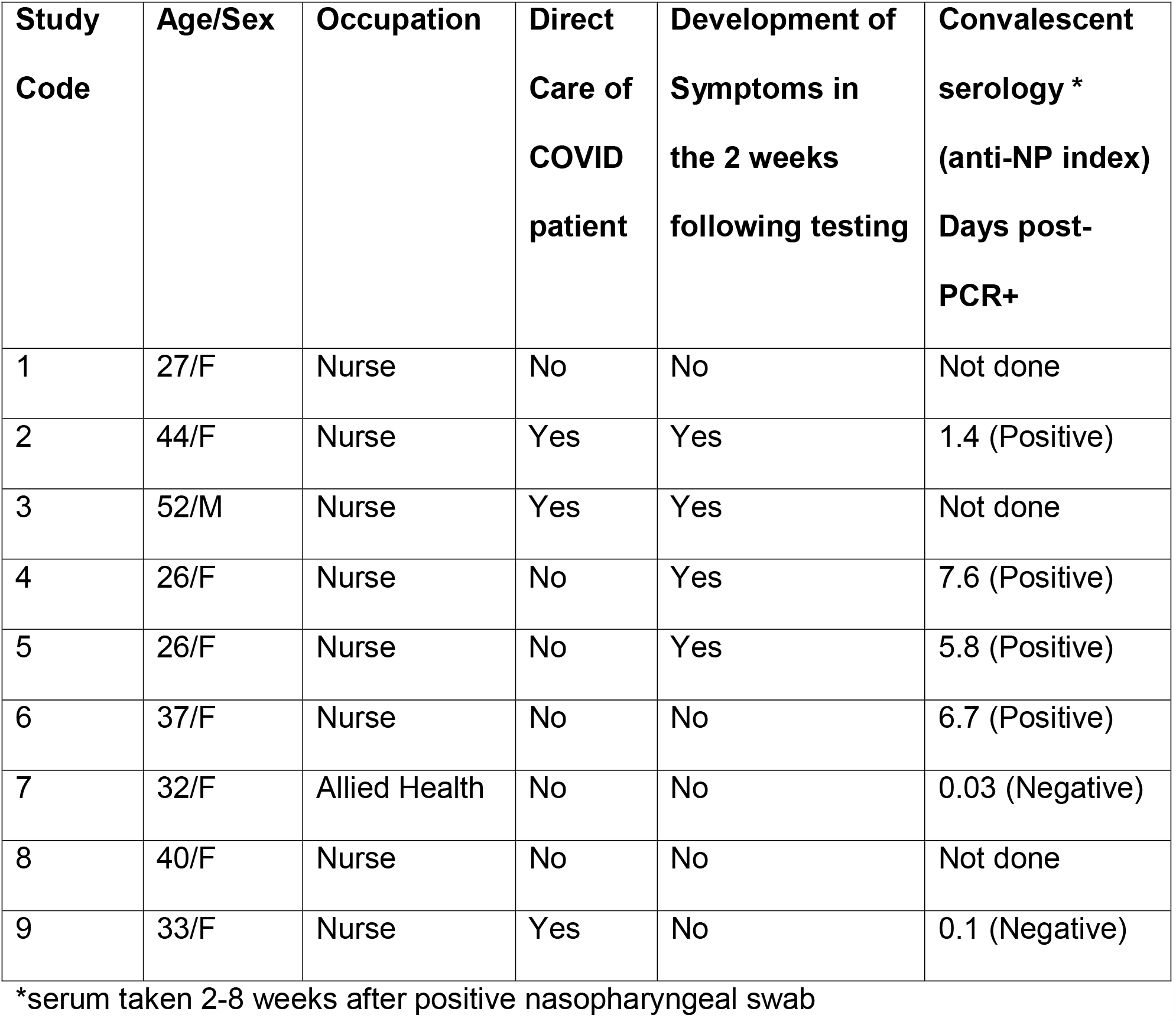
Asymptomatic Healthcare workers that had positive SARS-CoV-2 PCR (n=9) in Cohort 1.

| **Study Code** | **Age/Sex** | **Occupation** | **Direct Care of COVID patient** | **Development of Symptoms in the 2 weeks following testing** | **Convalescent serology \***<br>**(anti-NP index)**<br>**Days post-PCR+** |
| --- | --- | --- | --- | --- | --- |
| 1 | 27/F | Nurse | No | No | Not done |
| 2 | 44/F | Nurse | Yes | Yes | 1.4 (Positive) |
| 3 | 52/M | Nurse | Yes | Yes | Not done |
| 4 | 26/F | Nurse | No | Yes | 7.6 (Positive) |
| 5 | 26/F | Nurse | No | Yes | 5.8 (Positive) |
| 6 | 37/F | Nurse | No | No | 6.7 (Positive) |
| 7 | 32/F | Allied Health | No | No | 0.03 (Negative) |
| 8 | 40/F | Nurse | No | No | Not done |
| 9 | 33/F | Nurse | Yes | No | 0.1 (Negative) |

\*serum taken 2-8 weeks after positive nasopharyngeal swab

**Supplementary Table 4:** Healthcare Workers that were SARS-CoV-2 anti-nucleoprotein (NP) IgG positive (n=14).

| **Study Number** | **Age/Sex** | **Occupation** | **Direct Care of COVID patient** | **History of Recent Compatible Symptoms** | **SARS-CoV-2 PCR done on same day** |
| --- | --- | --- | --- | --- | --- |
| 1* | 27/F | Nurse | - | - | - |
| 2* | 45/F | Allied Health | + | - | - |
| 3 | 71/M | Physician | - | + | ND |
| 4 | 38/M | Nurse | - | - | - |
| 5 | 44/M | Nurse | - | + | - |
| 6 | 55/F | Nurse | - | - | - |
| 7 | 40/F | Allied Health | - | - | - |
| 8 | 61/F | Nurse | - | - | - |
| 9 | 46/F | Nurse | - | - | - |
| 10 | 39/F | Nurse | - | + | - |
| 11 | 36/F | Allied Health | - | - | - |
| 12 | 46/F | Allied Health | - | - | - |
| 13 | 58/F | Administrator | - | - | - |
| 14 | 36/M | Allied Health | - | + | - |

ND, not done
\*also anti-Spike IgG positive

**Supplementary Table 5:** List of antigen reactivities upregulated in COVID+ patients as determined by significance analysis of microarrays (fold change > 2, false discovery rate < 1%).

| **Antigen** | **Score** | **Fold Change** | **q-value (%)** |
| --- | --- | --- | --- |
| IgG_A43, SARS-CoV-2 (COVID-19) S1 Recombinant Protein | 7.75 | 71.38 | 0 |
| IgG_A31, SARS-CoV-2 (COVID-19, 2019-nCoV) S1+S2 ECD (S-ECD) Recombinant Protein | 7.36 | 36.60 | 0 |
| IgG_A2, SARS-CoV-2 (2019-nCoV) Spike Protein (RBD, His Tag) | 6.92 | 44.40 | 0 |
| IgG_A30, SARS-CoV-2 (COVID-19, 2019-nCoV) Spike RBD Recombinant Protein | 6.91 | 113.45 | 0 |
| IgG_A6, 2019-nCoV Spike Protein (S1 Subunit, His Tag) | 6.66 | 26.43 | 0 |
| IgG_A44, 2019-nCoV Nucleocapsid Recombinant Protein | 6.50 | 29.81 | 0 |
| IgG_A38, SARS-CoV-2 (COVID-19, 2019-nCoV) Spike-RBD Recombinant Protein | 6.12 | 79.78 | 0 |
| IgG_A42, 2019-nCoV Nucleocapsid Recombinant Protein | 5.94 | 43.87 | 0 |
| IgM_A30, SARS-CoV-2 (COVID-19, 2019-nCoV) Spike RBD Recombinant Protein | 5.18 | 12.15 | 0 |
| IgG_A26, SARS Coronavirus 2019 Spike Recombinant protein (1000-1200 aa) | 5.05 | 17.16 | 0 |
| IgG_A5, 2019-nCoV Spike Protein (S2 ECD, His tag) | 4.97 | 9.98 | 0 |
| IgG_A10, Human SARS Coronavirus Nucleoprotein / NP Protein (His Tag) | 4.53 | 17.43 | 0 |
| IgG_A29, SARS Coronavirus 2019 Spike E Mosaic Recombinant protein | 4.47 | 8.03 | 0 |
| IgG_A4, 2019-nCoV Nucleocapsid Protein (His tag) | 4.31 | 13.08 | 0 |
| IgM_A2, SARS-CoV-2 (2019-nCoV) Spike Protein (RBD, His Tag) | 4.29 | 6.03 | 0 |
| IgM_A43, SARS-CoV-2 (COVID-19) S1 Recombinant Protein | 4.25 | 8.07 | 0 |
| IgM_A6, 2019-nCoV Spike Protein (S1 Subunit, His Tag) | 3.99 | 7.44 | 0 |
| IgG_A16, MERS-CoV (NCoV / Novel coronavirus) Spike Protein (S2 Subunit, aa 726-1296, His Tag) | 3.81 | 5.54 | 0 |
| IgG_A37, SARS-CoV-2 (COVID-19, 2019-nCoV) Spike-ECD Recombinant Protein | 3.51 | 4.36 | 0 |
| IgG_A39, COVID 19 M Coronavirus Recombinant Protein | 3.27 | 2.62 | 0 |
| IgG_A3, 2019-nCoV Spike Protein (S1+S2 ECD, His tag) | 3.25 | 3.89 | 0 |
| IgG_A36, SARS-CoV-2 (COVID-19, 2019-nCoV) Spike-RBD Recombinant Protein | 2.99 | 6.01 | 0 |
| IgM_A31, SARS-CoV-2 (COVID-19, 2019-nCoV) S1+S2 ECD (S-ECD) Recombinant Protein | 2.92 | 2.82 | 0 |
| IgG_A24, MERS-CoV (NCoV / Novel coronavirus) Spike Protein (ECD, aa 1-1297, His Tag) | 2.90 | 3.24 | 0 |
| IgG_A35, SARS-CoV-2 (COVID-19, 2019-nCoV) Nucleocapsid Recombinant Protein | 2.57 | 2.78 | 0 |
| IgM_A42, 2019-nCoV Nucleocapsid Recombinant Protein | 2.16 | 2.41 | 0 |
| IgM_A23, Human coronavirus (HCoV-OC43) Hemagglutinin esterase Protein (His Tag) | 2.15 | 4.07 | 0 |
| IgM_A38, SARS-CoV-2 (COVID-19, 2019-nCoV) Spike-RBD Recombinant Protein | 2.14 | 3.78 | 0 |
| IgM_A25, SARS Coronavirus 2019 Spike Recombinant protein (800-1000 aa) | 2.12 | 2.98 | 0 |
| IgM_A15, MERS-CoV (NCoV / Novel coronavirus) Spike Protein (S1 Subunit, aa 1-725, His Tag) | 1.99 | 2.67 | 0 |
| IgM_A44, 2019-nCoV Nucleocapsid Recombinant Protein | 1.99 | 2.11 | 0 |
| IgM_A26, SARS Coronavirus 2019 Spike Recombinant protein (1000-1200 aa) | 1.75 | 2.43 | 0 |
| IgM_A22, Human coronavirus (HCoV-NL63) Spike/S1 Protein (S1 Subunit, His Tag) | 1.75 | 2.64 | 0 |
| IgM_A7, SARS-CoV-2/2019-nCoV Plpro / papainlike protease (aa 1564-1880, His Tag) | 1.71 | 2.34 | 0 |
| IgM_A5, 2019-nCoV Spike Protein (S2 ECD, His tag) | 1.61 | 2.34 | 0 |
| IgG_A11, Human SARS Coronavirus Spike Protein (S1 Subunit, His Tag) | 1.58 | 2.47 | 0 |
| IgM_A19, Human coronavirus spike glycoprotein Protein (isolate HKU1) (S1 Subunit, aa 1-760, His Tag) | 1.48 | 2.41 | 0 |
| IgM_A3, 2019-nCoV Spike Protein (S1+S2 ECD, His tag) | 1.47 | 2.12 | 0 |
| IgM_A41, SARS-CoV-2 (COVID-19) Papain-like Protease | 1.46 | 2.20 | 0 |

**SUPPLEMENTARY FIGURE 1:**
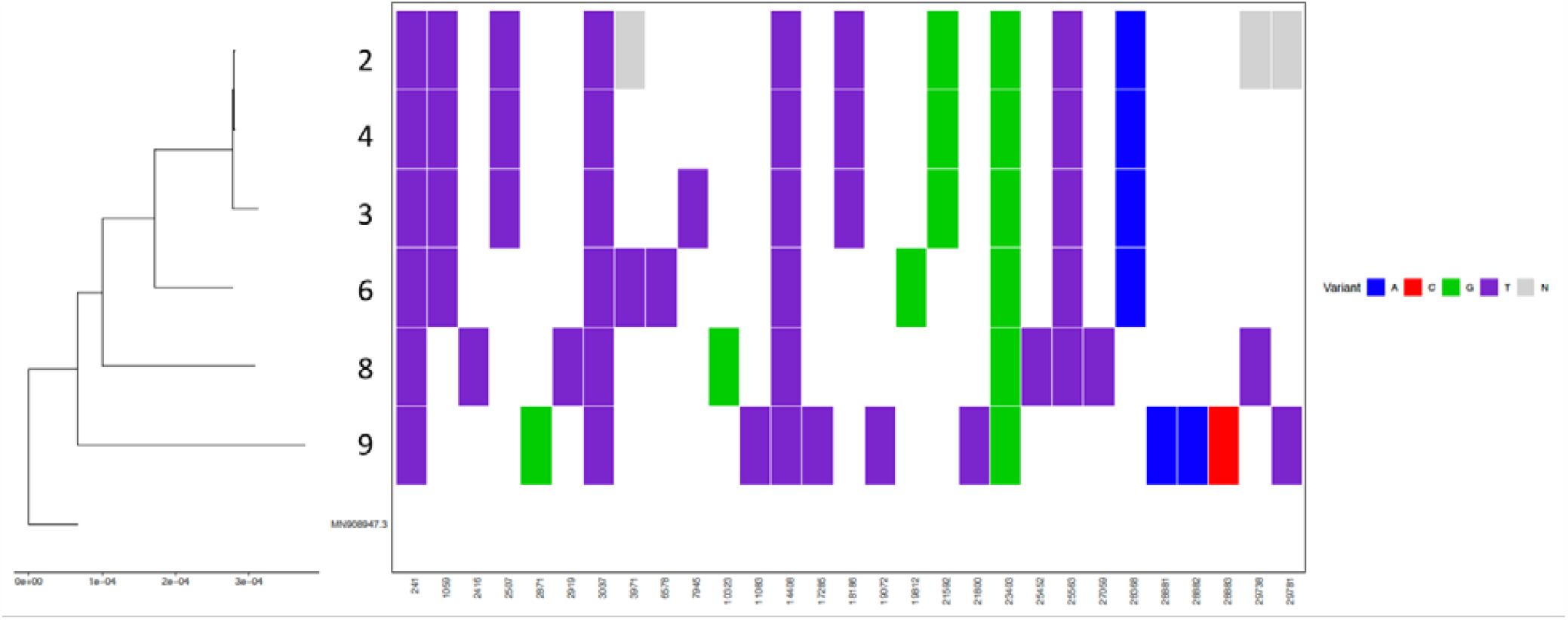
Sequencing of SARS-CoV-2 from 6/9 healthcare workers with active infection (numbers on y-axis correspond to those in Supplementary Table 3). A multiple sequence alignment of all consensus reads and the MN908947.3 reference was generated, then used to build a phylogenetic tree using augur (https://github.com/nextstrain/augur). Variants were called using scripts developed as part of the nCoV-tools package (https://github.com/jts/ncov-tools). Sites with single base substitutions are shown, with N indicating no coverage at the site. For genome completeness, a cut-off of 75% was used to sequence samples. Three of the nine samples did not meet this cut-off. Genome completeness ranged between 83.7-97.1%. Results demonstrate 3 variants.

**SUPPLEMENTARY FIGURE 2:**
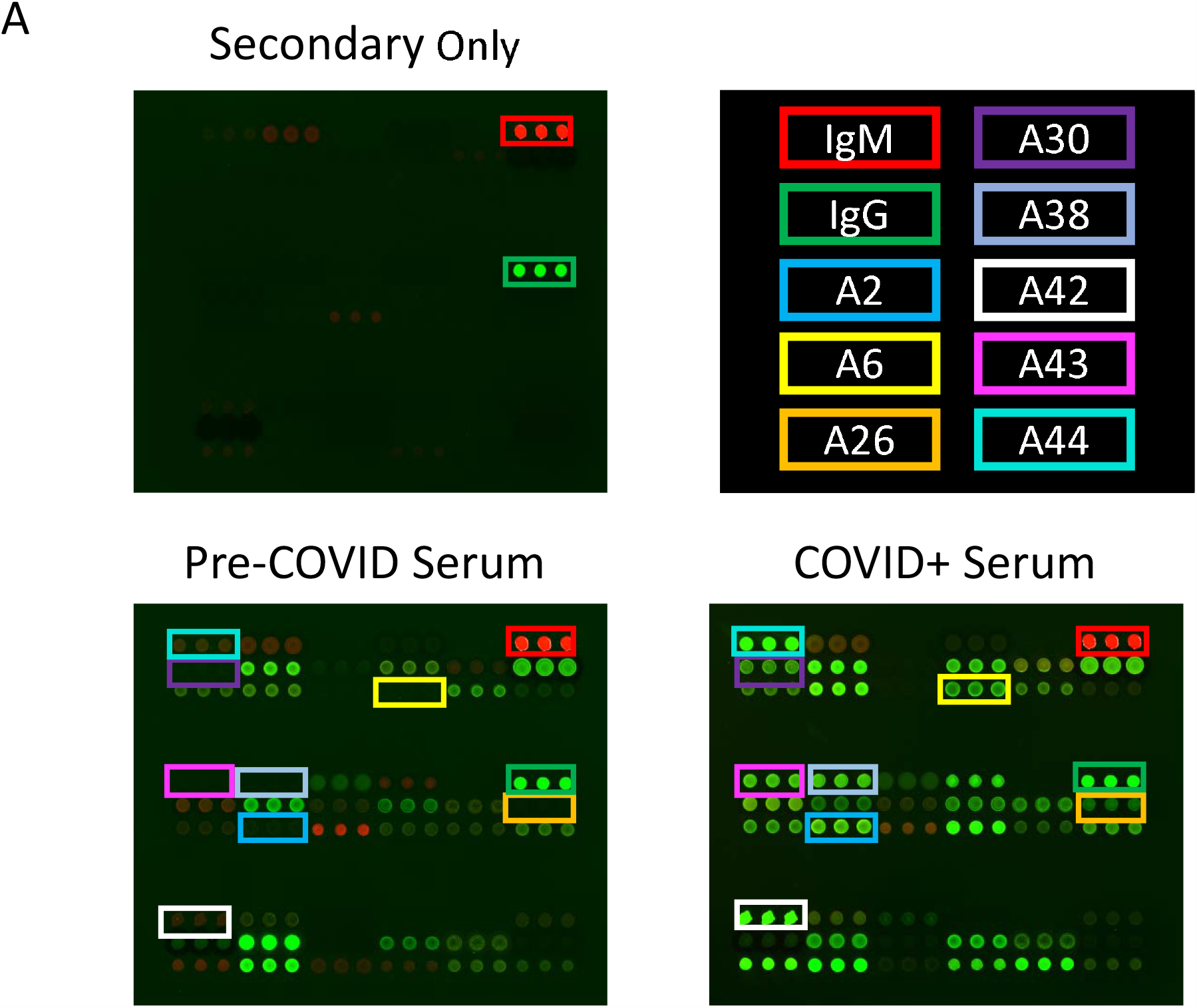
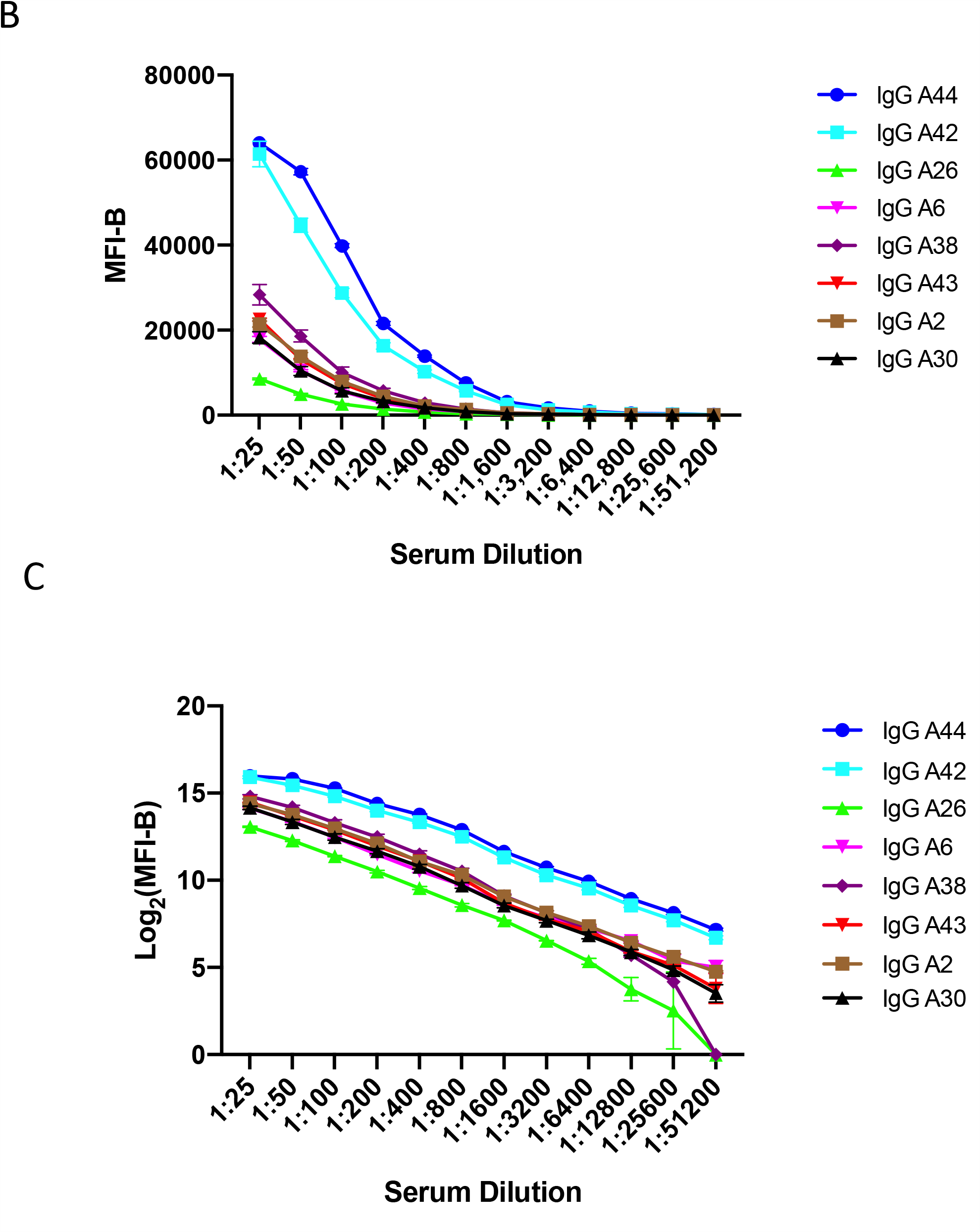
Images of antigen microarrays and determination of linearity of the array assay. A) Images of 2-color arrays probed with secondary antibodies only, pre-COVID serum (negative control) and COVID+ serum (positive control). Antigens were spotted in triplicate; green indicates IgG reactivity, whereas red indicates IgM reactivity. On the array probed only with secondary antibodies, only human IgG and human IgM are detected. On the array probed with pre-COVID serum, reactivity against common community coronavirus antigens is detected. On the array probed with COVID+ serum, there are additional SARS-CoV-2 reactivities detected (boxes). Array features are approximately 500 μm in diameter. B) and C) Linearity studies using serial dilutions of COVID+ serum. Graph B shows MFI-B plotted against serum dilutions, whereas Graph C shows log_2_ transformed MFI-B. Linear responses are observed over a wide range of serum dilutions using log_2_ transformed MFI-B. Antibody responses become non-linear as MFI-B approaches saturation levels (MFI-B > 60,000). MFI-B, median fluorescent intensity minus background.

**SUPPLEMENTARY FIGURE 3:**
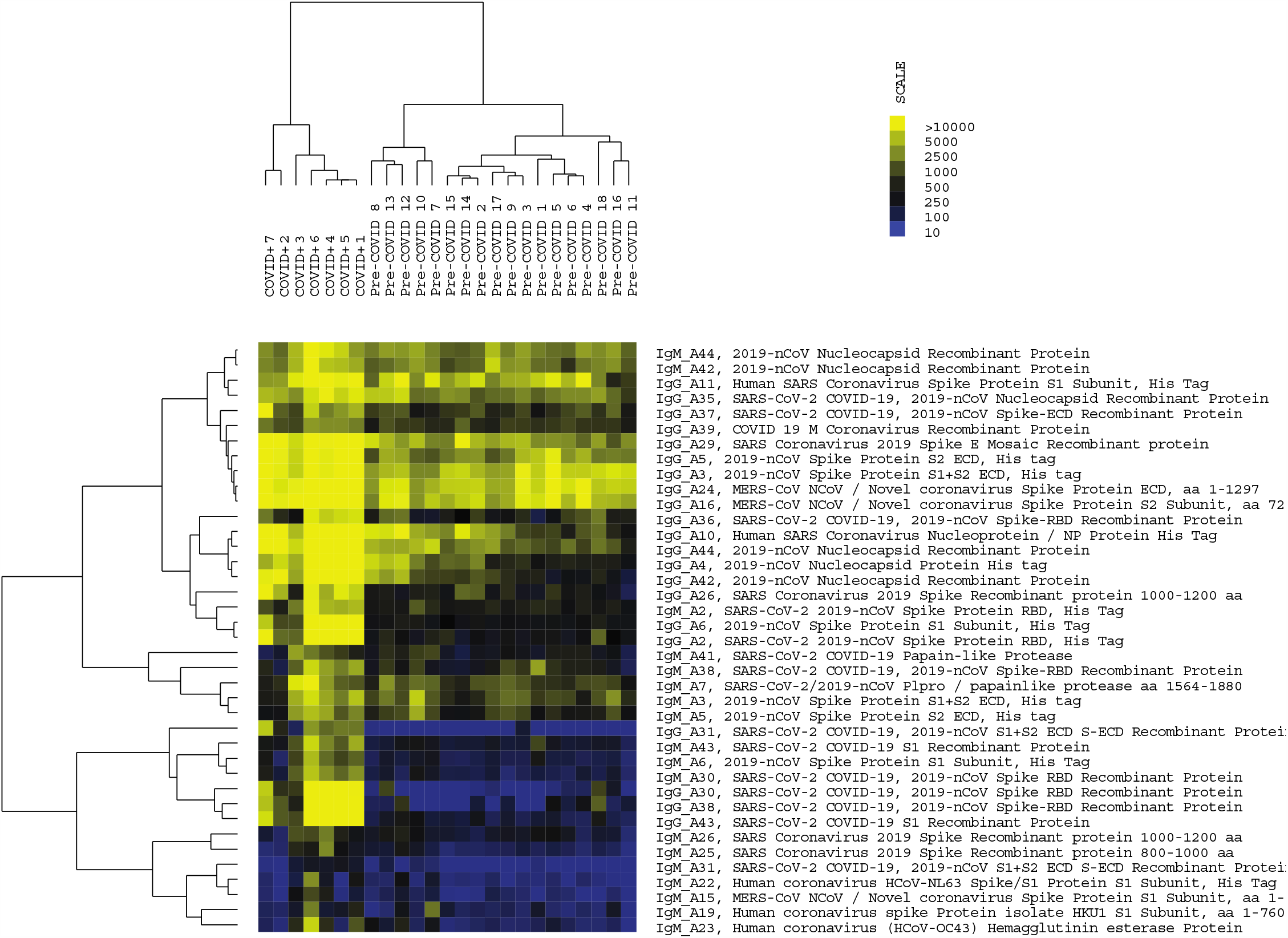
Heatmap of the 39 antigen reactivities upregulated in COVID+ patients as determined by significance analysis of microarrays. The COVID+ samples (n=7) form a separate cluster from the pre-COVID samples (n=18) using a hierarchical clustering algorithm. Yellow indicates high reactivity, whereas blue indicates low reactivity.

## Notes

### Competing Interest Statement

The authors have declared no competing interest.

### Clinical Trial

This was not a clinical trial. No interventions took place.

### Funding Statement

The study was funded by the Mount Sinai Hospital and University Health Network Academic Medical Organization. It was also funded by the Toronto General and Western Foundation. No author or institution received payment or services from a third party for any aspect of the submitted work.

### Author Declarations

The study was approved by the University Health Network Research Ethics Board.

